# Effectiveness of a Relaxation Technique on Reducing Anxiety, Depression, and Fatigue among Women with Breast Cancer in Goa: A Randomized Controlled Trial

**DOI:** 10.64898/2025.12.17.25342348

**Authors:** Dhanya Jose, Jagdish Anil Cacodcar

## Abstract

**Objective:** Psychological distress in cancer is associated with poorer quality of life, poorer treatment adherence and outcomes, and higher healthcare costs. The current study aimed to provide evidence on the effectiveness of the Progressive Muscle Relaxation program in female patients with breast cancer at Goa Medical College, Goa, a state in the Western part of India. This study will help to explore the level of anxiety, depression, and fatigue and provide suitable recommendations to reduce anxiety, depression, and fatigue among cancer patients.

**Methods:** It was a Randomised Controlled Trial. The study was conducted in Surgery Wards 106 and 109 at Goa Medical College. Adult women ≥ 18 years of age with a new diagnosis of breast cancer who have undergone surgery as their primary mode of treatment are included in the study. Sixty patients were randomly divided into two groups of 30 each, using a block randomization method with ten numbered sealed opaque envelopes. Fifty-eight people completed the study. The intervention entailed a 20-minute Progressive Muscle Relaxation (PMR) session, which started with Deep Breathing and Guided Imagery (GI) sessions given to one group. Both groups were evaluated for anxiety, depression, and fatigue levels using the Zung Anxiety Scale, Beck Depression Inventory, and Fatigue Scale at baseline and after the two-week intervention.

**Results:** Fifty-eight people completed the study. The participants in both groups had similar sociodemographic characteristics and clinical profiles. After two weeks of intervention, the intervention group showed significant reductions in anxiety (51.71 ± 2.89 to 38.52 ± 6.32, p ≤ 0.001), depression (11.48 ± 2.93 to 6.16 ± 2.98, p ≤ 0.001), and fatigue (23.29 ± 4.12 to 16.88 ± 4.73, p ≤ 0.001).

**Conclusions:** This study highlights the effectiveness of Progressive Muscle Relaxation (PMR) and Guided Imagery (GI) in reducing anxiety, depression, and fatigue in breast cancer patients. These nonpharmacological techniques serve as valuable complementary therapies, helping manage emotional distress and prevent symptom progression during treatment.

**Trial registration number and date:** CTRI (Clinical Trial Registry of India) Registration was done in February 2021 from the Clinical Trial Registry of India, with Reg. number CTRI/2021/02/030996.

## 1 Introduction

Cancer is a significant cause of premature death in 134 out of 183 countries. In 2020, there were 2.3 million women diagnosed with breast cancer and 6,85,000 deaths globally. As of the end of 2020, 7.8 million women have been diagnosed with breast cancer who are alive in the past five years, making breast cancer the most prevalent cancer in the world [1]. As per the GLOBOCAN data 2020, in India, Breast Cancer accounted for 13.5% (1,78,361/out of 1324413) of all cancer cases and 10.6% of all deaths. The frequency of breast cancer is 16.84% in the sample population from Goa [2].

The prevalence of anxiety and depression among patients with malignancy is up to 49% [3]. Psychological distress in cancer is associated with poorer quality of life, poorer treatment adherence and outcomes, and higher healthcare costs. It can have a detrimental effect on overall survival [4]. Identifying cancer patients who may be more likely to suffer from psychological distress over the disease trajectory is essential to targeting the proper interventions and providing the best care [4]. Cancer-related fatigue (CRF) is a frequently reported and distressing symptom. It is often more debilitating than pain and significantly impacts the quality of life (QOL) of cancer patients. [5] Fatigue can also be part of depression. Preventing or reducing mental health problems amongst cancer patients is an essential public health and clinical concern.

To manage anxiety, depression, and fatigue, patients often turn to medication, psychotherapy, and cognitive behavioral techniques. Studies have found that psychological techniques can help patients feel more in control of uncomfortable symptoms and cancer treatment side effects. These techniques include simple thought reframing, deep breathing, art and music therapy, progressive muscle relaxation (PMR), and guided imagery (GI) [6]. When a person is anxious, there are three different components to their reaction. The physiological component is increased heart rate, sweating, and muscle tension; the behavioral component like avoidance, trying to escape, and the cognitive component is negative thoughts such as I am going to collapse, and I cannot cope. One effective way of breaking this vicious circle is to focus on the physiological reaction and learn how to control it. As anxiety can build up very quickly and occur in a wide range of situations, effective relaxation would allow individuals to relax not just when sitting in a chair at home but in any situation and do so very quickly. This is the aim of applied relaxation training [7]. PMR, which stands for “Progressive Muscle Relaxation,” is considered one of the most common and widely used applied relaxation training techniques, where individuals systematically tense and then release different muscle groups in the body to achieve a state of deep relaxation and reduce stress or anxiety. It is often referred to as “JPMR,” which stands for “Jacobson’s Progressive Muscle Relaxation” - named after the developer of the technique, Dr. Edmund Jacobson [8].

There are various studies done in different parts of the world regarding the effectiveness of progressive muscle relaxation techniques in reducing anxiety and depression in breast cancer and other cancer patients [6,9-12]. Recently, a review of eight randomized control trials among cancer patients has been published, which reported a positive impact on the mental state as well as on chemotherapy-induced nausea and vomiting [13]. One study from the Post Graduate Institute, Chandigarh, on cervical cancer patients undergoing chemotherapy was the only Indian study identified in this regard [6]. The current study aimed to provide evidence on the effectiveness of the Jacobson Progressive Muscle Relaxation (PMR) technique in breast cancer females at Goa Medical College, Goa, a State in the Western part of India. This hospital is the only tertiary care facility in the State and caters to patients from all over the State. No study has been published yet on this vital topic in Goa. This study will help to explore the level of anxiety and depression and provide suitable recommendations to reduce anxiety and depression among cancer patients.

### 1.1 Objectives of the study

a. Examine the effectiveness of a relaxation technique in reducing anxiety, depression, and fatigue in women with breast cancer.

b. Estimate the level of anxiety, depression, and fatigue among women with breast cancer.

## 2 Materials & Methods

A Randomized Controlled Trial was conducted from February 2021 to May 2022. It was a Parallel Group Trial, hospital-based study in which participants were recruited from Surgery wards 106 and 109 at Goa Medical College, Goa, India. The inclusion criteria were the following: (1) Adult women ≥ 18 years of age with a new diagnosis of breast cancer who have undergone surgery (mastectomy) as their primary mode of treatment. (2) Those who experienced anxiety or depression (Zung anxiety scale score; SAS > 44, Beck Depression Inventory; BDI > 10, assessed by the principal investigator, details mentioned in outcome assessment section) were not receiving medication for anxiety or depression. (3) Those who do not have cognitive impairment and (4) those who consent to participate in the study. Patients who required psychiatry consultation or medication for anxiety or depression during the study period (SAS > 60, BDI > 17) were excluded from the study.

### 2.1 Sample size

The sample size N was calculated based on inputs from similar prior studies with 95% confidence and 90% power.

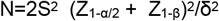

Here, Z_1-α/2_ = 1.96, Z_1-β_= 1.28, S^2^ = sample variance (S.D) ^2^, and δ = difference in anxiety score [14]. From the study by Kaur et al., the posttest anxiety scores 32.23±3.22 and 39.77±7.55 were taken for the calculation. The required sample size is computed as N = 2 (1.96 + 1.28 / 7.54)^2^ 7.55^2^ ≈ 21, that is 21 per group. Considering the 15% loss to follow-up, we took a sample size of 30 per group [6].

### 2.2 Trial Registration

This study was performed in line with the principles of the Declaration of Helsinki. Ethical approval was obtained in December 2020 from the Institutional Ethics Committee (IEC) of Goa Medical College. The amendment was made on 13/05/2022 regarding restricting the study to breast cancer, which was initially planned to include other gynecological cancers, too, considering the sample adequacy during the COVID-19 epidemic. CTRI (Clinical Trial Registry of India) Registration was done in February 2021 from the Clinical Trial Registry of India, with Reg. number CTRI/ 2021/02/030996, where the full trial protocol can be accessed [15]. The RCT has been reported following the CONSORT guidelines.

### 2.3 Funding

Financial support and sponsorship / Conflicts of interest are not declared.

We obtained written informed consent from each participant. All data collected from participants was kept strictly confidential, and apart from the study investigators, no information was given to anyone, including institutional authorities. The participants’ names and identifying information will not be disclosed in any publication.

### 2.4 Intervention and Procedures

The patients were provided with information sheets during the postoperative period and invited to participate in the study. Patients willing to participate in the study were assessed for eligibility. Out of 104 people who were invited to the study, 44 were excluded. One declined to participate, 2 were done only lumpectomy not mastectomy, 41 did not have the Zung anxiety scale score; SAS > 44, Beck Depression Inventory; BDI > 10. Nobody was excluded due to high scores (SAS ≥ 60, BDI ≥ 17), which required psychiatry consultation or medication for anxiety or depression. (Figure 1)

**Figure 1.**
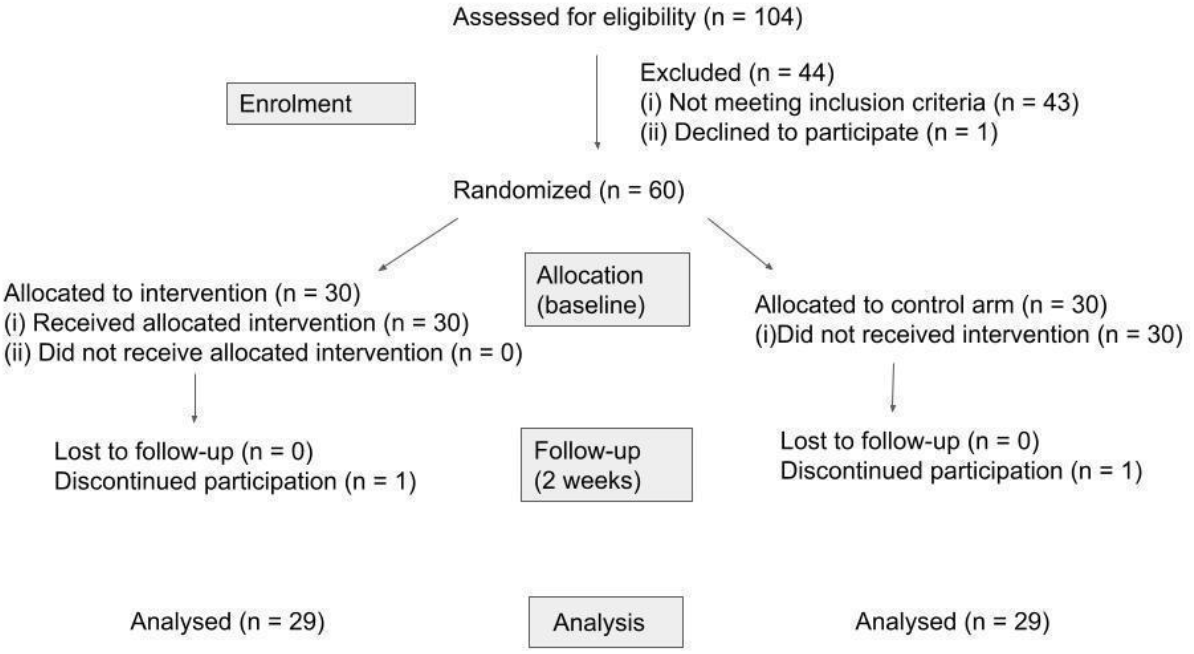
CONSORT flow diagram

#### 2.4.1 Randomization and Masking

After getting consent, baseline questionnaires were completed before randomization. Consenting patients were randomly assigned in a 1:1 ratio to intervention or control groups by selecting from a block of ten numbered, sealed opaque envelopes. The method of Generating Random Sequence was Computer-generated randomization. Method of Concealment was sequentially numbered, sealed, opaque envelopes. This was an open-blinded clinical trial. The blinding was impractical since the intervention was an exercise technique [16].

#### 2.4.2 Intervention

If the patient was allocated to the intervention group, the intervention session took place the next day. The intervention entailed a 20-minute relaxation session, a combination of Progressive Muscle Relaxation (PMR), Deep Breathing, and Guided Imagery (GI) sessions. Progressive Muscle Relaxation (PMR) is a method that involves systematically tensing and then relaxing different muscle groups throughout the body to promote physical and mental relaxation. Guided Imagery (GI) is a cognitive technique that uses imagination to create positive mind-body responses and activate the senses [9, 17, 18]. Deep breathing consists of breathing through the belly and not the chest, more slowly and deeply. Deep breathing lowers your heart rate and your blood pressure increases your energy level and decreases muscle tension and pain. The relaxation technique was taught by the researcher only. Patients were encouraged to ask questions about the intervention. The interview was conducted in a quiet area in the ward, and every effort was taken to avoid interruptions during the session. Patients in the control and intervention groups were provided with all standard post-operative support provided by the Goa Medical College.

The homework assignment was to practice progressive relaxation for approximately 15-20 minutes twice a day and whenever they feel anxious or stressed. Patients were requested to do it in a place and time where they would be comfortable and unlikely to be interrupted. They were also requested to keep a record of the time taken to relax and the amount of relaxation achieved (0-100 scales) during each practice. To ensure consistency in the exercises, an instruction sheet was provided. Daily reminders (text messages) were given.

Two weeks following surgery, at the time of the first review, subjects completed the follow-up questionnaires. Patients who received the intervention were asked to provide feedback about the session. Patients who did not receive the intervention were taught the relaxation technique during this visit. Hence, the control group can be described as a wait-list control, as participants in the control arm received the PMR + GI intervention after the end of the study period.

### 2.5 Outcome Assessment

Both groups were assessed for anxiety and depression levels at baseline and after the two-week intervention. Anxiety was measured using the Zung Self-Rating Anxiety Scale (SAS), a 20-item self-report questionnaire evaluating the presence and severity of anxiety symptoms. Depression was assessed with the Beck Depression Inventory-II (BDI-II), a 21-item tool designed to measure common depressive symptoms [9, 19, 20]. The Fatigue Assessment Scale (FAS) was also used to measure fatigue levels [9, 21]. Patients with scores of SAS > 60 and BDI > 17 were excluded from the study as they required psychiatry consultation for anxiety or depression during the study period.

Sociodemographic data and clinical case details were collected. Vital signs were recorded. Patients’ feedback about the relaxation techniques was collected at the end of the study. The primary outcome was the difference between baseline and follow-up scores on anxiety, depression, and fatigue scales. Secondary outcomes included the scores of anxiety, depression, and fatigue scales and the patient’s overall impression of the intervention.

### 2.6 Data analysis

The data was entered using Google Sheets and analyzed using R-program software. p-value < 0.05 was taken as a level of significance. Student t-test and χ 2 test were used for statistical analysis. For a given scale, mean changes from baseline to the end of the intervention were analyzed using a paired t-test, while mean differences between the two groups were assessed with an unpaired t-test.

## 3 Results

Between February 2021 and May 2022, 95 patients were evaluated for eligibility, 60 were enrolled, and 58 successfully completed all assessments. The final sample comprised 58 female breast cancer patients, evenly distributed between the intervention and control groups, with 29 participants in each. At baseline, both groups were generally comparable in key demographic variables.

### 3.1 Sociodemographic profile of the study participants (n=58)

Table 1 presents the sociodemographic profile of the 58 patients included in the study. Both groups were comparable in terms of age, religion, habitat, marital status, education level, occupation, and per capita income. The mean age was 57.90 ± 12.70 years (range: 30-81) in the experimental group and 57.66 ± 13.03 years (range: 30-81) in the control group.

**Table 1.**
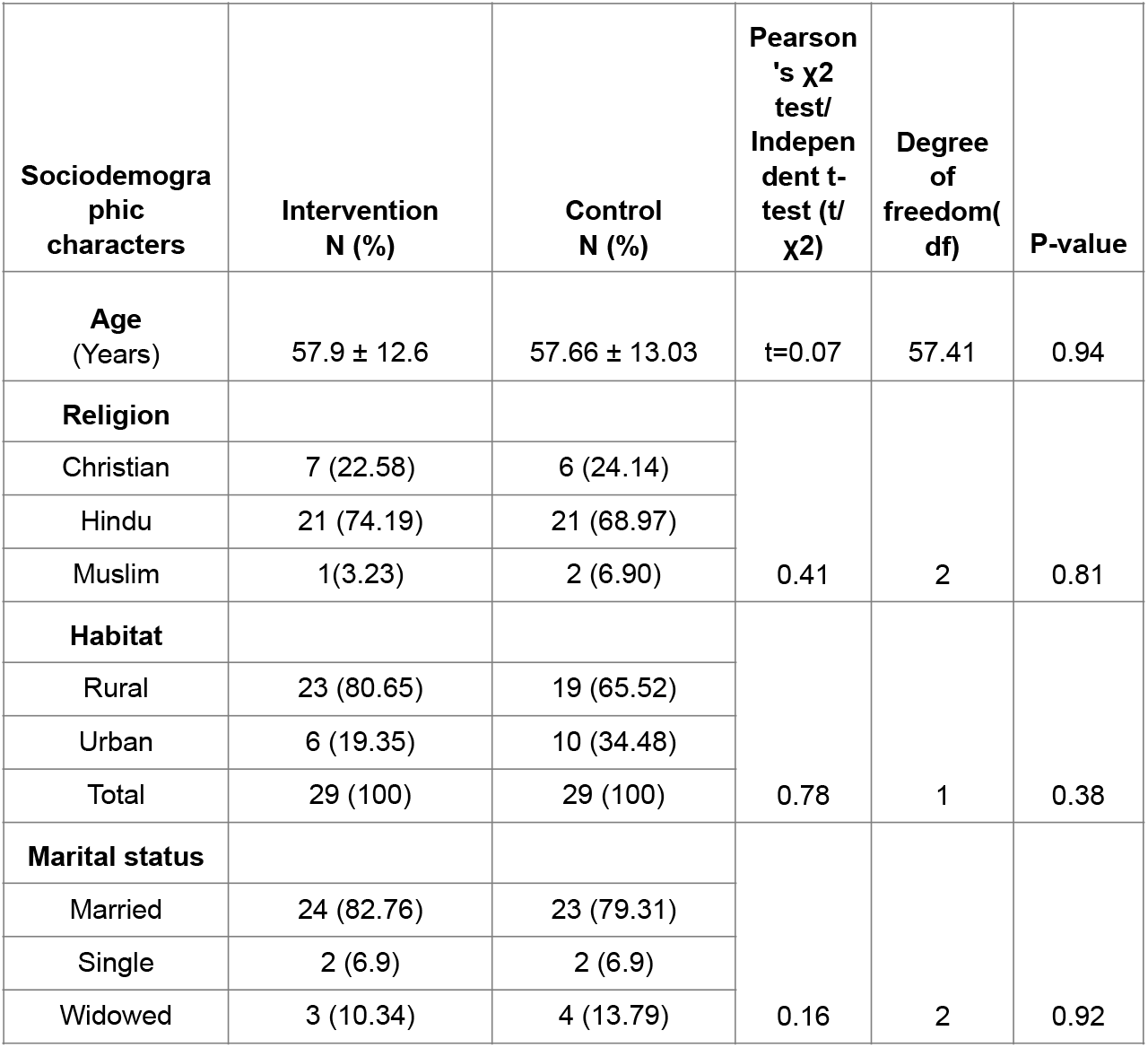
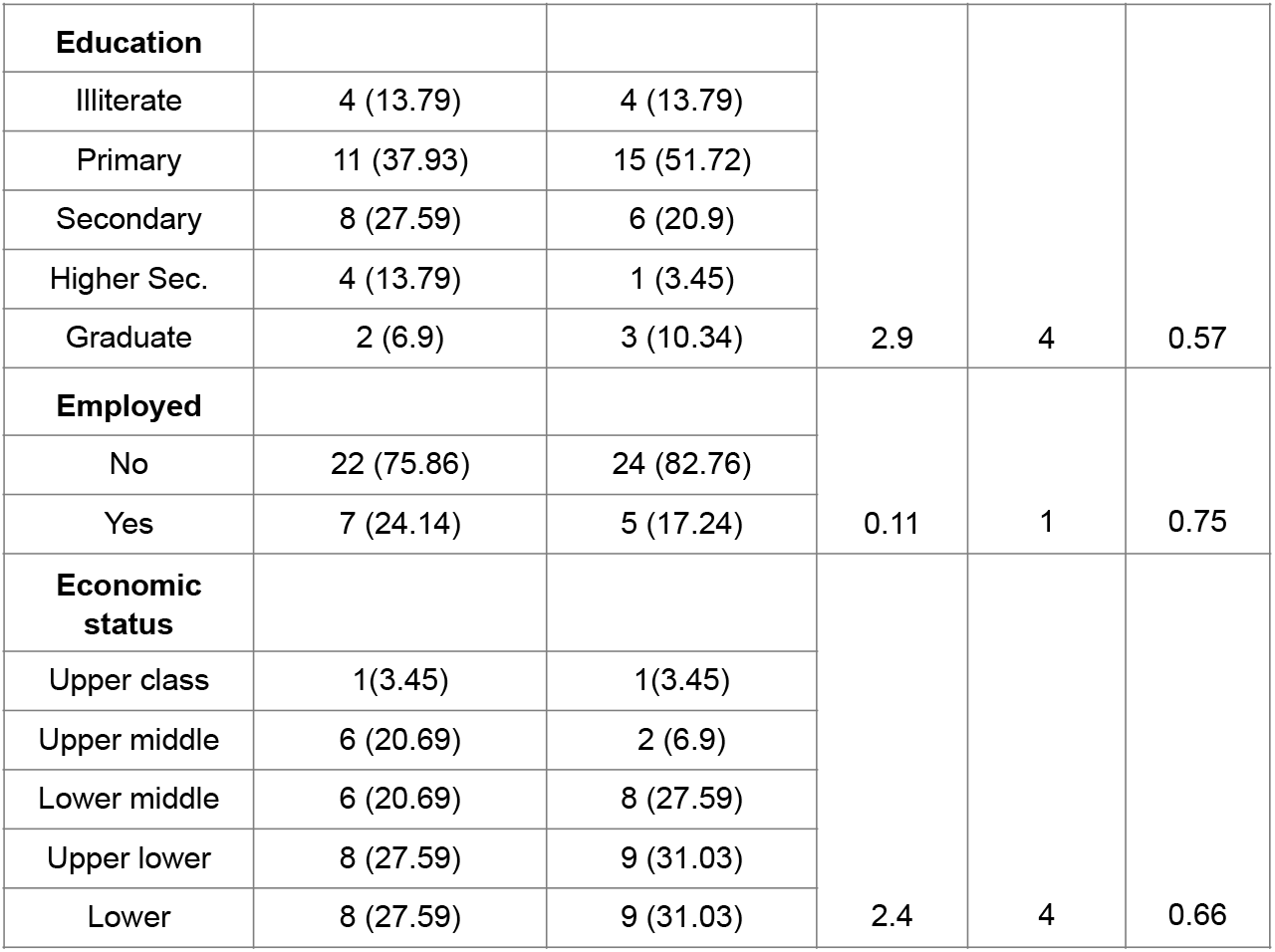
Sociodemographic profile of the study participants (n=58)

The majority of patients in the experimental group (21, 74.19%) and more than half in the control group (21, 68.97%) were Hindu. A higher proportion of patients in the experimental group (23, 80.65%) compared to the control group (19, 65.52%) resided in rural areas. Most participants were married, with 24 (82.76%) in the experimental group and 23 (79.31%) in the control group.

Regarding education, most patients in both groups had primary or secondary education. In terms of employment, 22 (75.86%) patients in the experimental group and 24 (82.76%) in the control group were unemployed.

More than half of the patients in the experimental group and the majority in the control group had a per capita monthly income of less than ₹3,765 and fell into the lower middle, upper lower, or lower socioeconomic classes based on B.G. Prasad’s socioeconomic scale.

### 3.2 Clinical profile of the breast cancer patients (n = 58)

Table 2 presents the clinical profile of patients in both the experimental and control groups. Overall, the clinical characteristics were comparable between the two groups. More than half of the patients had an ASA score of 2, with 16 (55.17%) in the experimental group and 17 (58.62%) in the control group. (The American Society of Anesthesiology score is a metric to determine if someone is healthy enough to tolerate surgery and anesthesia). The mean length of surgery was 177.76 minutes in the intervention group and 165.1 minutes in the control group. Postoperative hospital stay was 11.55 and 12.52 days in both the intervention and control groups, respectively. For the remaining few days, an average of 2 days, they did exercise from home. The mean symptom duration length before the medical presentation was 155.38 and 109.03 days, respectively, in the intervention and control groups. The time duration from medical presentation to treatment in both the experimental and control groups was around three weeks. All patients underwent modified radical mastectomy as the primary mode of management. Most of our patients were under the histological classification of invasive ductal carcinoma (IDC); the remaining cases are classified as invasive lobular carcinoma (ILC). Most of them came under IDC Grade 2, 2-3, and 3 in both groups. According to AJCC anatomic staging, all our patients were between stage 1 and stage 3 B. Most of them were in Stage 2B in both the intervention and control groups, as seen in Table 2. More than half of the patients in both the intervention and control groups were classified under BI-RADS 5 in the radiologic staging of breast cancer. The majority of patients, 27 (93.1%) in both the experimental and control groups, presented with symptoms of a lump in the breast. Except for two patients in both the experimental and control groups, all had a history of breastfeeding. A family history of breast cancer was absent in the intervention group. Only 2 (6.9%) patients had a positive family history of breast cancer in the control group. Blood pressure and heart rate were also comparable in both groups.

**Table 2.** Clinical profile of the breast cancer patients (n = 58) *ASA-American Society of Anesthesiologists, IDC-Invasive Ductal Carcinoma, ILC-Invasive Lobular Carcinoma, BIRADS-Breast Imaging-Reporting and Data System, BP-Blood Pressure*

| Variable | Intervention<br>N (%) | Control<br>N (%) | Pearson's $\chi^2$ test/<br>Independent t-test<br>(t/$\chi^2$) | Degree<br>of<br>freedom (df) | P-value |
| --- | --- | --- | --- | --- | --- |
| ASA score |  |  |  |  |  |
| 1 | 8 (27.59) | 9 (31.03) | $\chi^2 = 0.59$ | 2 | 0.74 |
| 2 | 16 (55.17) | 17 (58.62) |  |  |  |
| 3 | 5 (17.24) | 3 (10.34) |  |  |  |
| Length of surgery (min) |  |  |  |  |  |
| Mean | 177.76±50.38 | 165.1±38.66 | t = 1.07 | 52.49 | 0.29 |
| Postoperative hospital stay (days) |  |  |  |  |  |
| Mean | 11.55±2.97 | 12.52±5.08 | t = -0.88 | 45.14 | 0.38 |
| Symptom duration (days) |  |  |  |  |  |
| Mean | 155.38±277.9<br>8 | 109.03±127.6<br>7 | t = 0.96 | 43 | 0.34 |
| Time from medical presentation to treatment (days) |  |  |  |  |  |
| Mean | 20.83±4.14 | 21.48±6.19 | t = -0.40 | 55.99 | 0.69 |
| Tumor type and grade |  |  |  |  |  |
| IDC Grade 2 | 12 (41.38) | 8 (27.59) |  |  |  |
| IDC Grade 2-3 | 10 (34.48) | 14 (48.28) |  |  |  |
| IDC Grade 2-3<br>& ILC | 1 (3.45) | 0 |  |  |  |
| IDC Grade 3 | 4 (13.79) | 2 (6.9) |  |  |  |
| IDC Grade 4 | 1 (3.45) | 0 |  |  |  |
| IDC with<br>sarcomatoid<br>differentiation<br>(Metaplastic<br>Ca.) | 0 | 2 (6.9) |  |  |  |
| ILC | 0 | 2 (6.9) |  |  |  |
| ILC Grade 3 &<br>IDC | 1 (3.45) | 0 |  |  |  |
| Mucinous carcinoma | 0 | 1 (3.45) | $\chi^2 = 10.13$ | 8 | 0.26 |
| Stage of malignancy |  |  |  |  |  |
| Stage 1 | 0 | 1 (4.76) | $\chi^2 = 5.21$ | 4 | 0.27 |
| Stage 2A | 4 | 5 (4.76) |  |  |  |
| Stage 2B | 16 (70.83) | 17 (76.19) |  |  |  |
| Stage 3A | 6 (12.5) | 1 (4.76) |  |  |  |
| Stage 3B | 3 (16.67) | 5 (9.52) |  |  |  |
| BIRADS |  |  |  |  |  |
| BIRADS 3 | 1 (3.45) | 0 | $\chi^2 = 5.18$ | 5 | 0.39 |
| BIRADS 4a | 0 | 2 (7.14) |  |  |  |
| BIRADS 4b | 3 (10.34) | 2 (7.14) |  |  |  |
| BIRADS 4c | 1 (3.45) | 3 (10.71) |  |  |  |
| BIRADS 5 | 18 (62.07) | 18 (64.29) |  |  |  |
| BIRADS 6 | 6 (20.69) | 3 (10.71) |  |  |  |
| Presenting symptom |  |  |  |  |  |
| Lump | 28 (96.55) | 28 (96.55) | $\chi^2 = 0$ | 1 | 1 |
| Ulceration | 1 (3.45) | 1(3.45) |  |  |  |
| Breastfeeding history |  |  |  |  |  |
| No | 2 (6.9) | 2 (6.9) | $\chi^2 = 0$ | 1 | 1 |
| Yes | 27 (93.1) | 27 (93.1) |  |  |  |
| Family history of breast cancer |  |  |  |  |  |
| No | 29 (100) | 27 (93.1) | $\chi^2 = 0.52$ | 1 | .47 |
| Yes | 0 | 2 (6.9) |  |  |  |
| Systolic BP |  |  |  |  |  |
| Mean | 120.83±10.97 | 119.79±12.12 | t = 0.34 | 55.45 | 0.73 |
| Diastolic BP |  |  |  |  |  |
| Mean | 77.1±6.54 | 76.38 ±7.24 | t = 0.4 | 55.42 | 0.69 |
| Heart rate |  |  |  |  |  |
| Mean | 83.07±6.49 | 80.9±7.26 | t = 1.22 | 54.14 | 0.23 |

### 3.3 Measurement of baseline level of anxiety, depression, and fatigue in breast cancer patients (N=99)

Measurement of baseline anxiety showed minimal to moderate levels of anxiety in 29 (100%) in both the intervention and control groups, according to the Zung Self-Rating Anxiety Scale (SAS). Measurement of baseline depression according to the Beck Depression Inventory (BDI) showed mild mood disturbance in the majority of patients from both intervention, 22 (75.86%) and control, 19 (65.52%) group. Measurement of baseline fatigue according to the Fatigue Assessment Scale (FAS) showed moderate level fatigue in 22 (75.86%) patients from the intervention group and 21 (72.41%) patients from the control group. Both groups were comparable with a baseline measurement of anxiety, depression, and fatigue. (See Table 3)

**Table 3.**
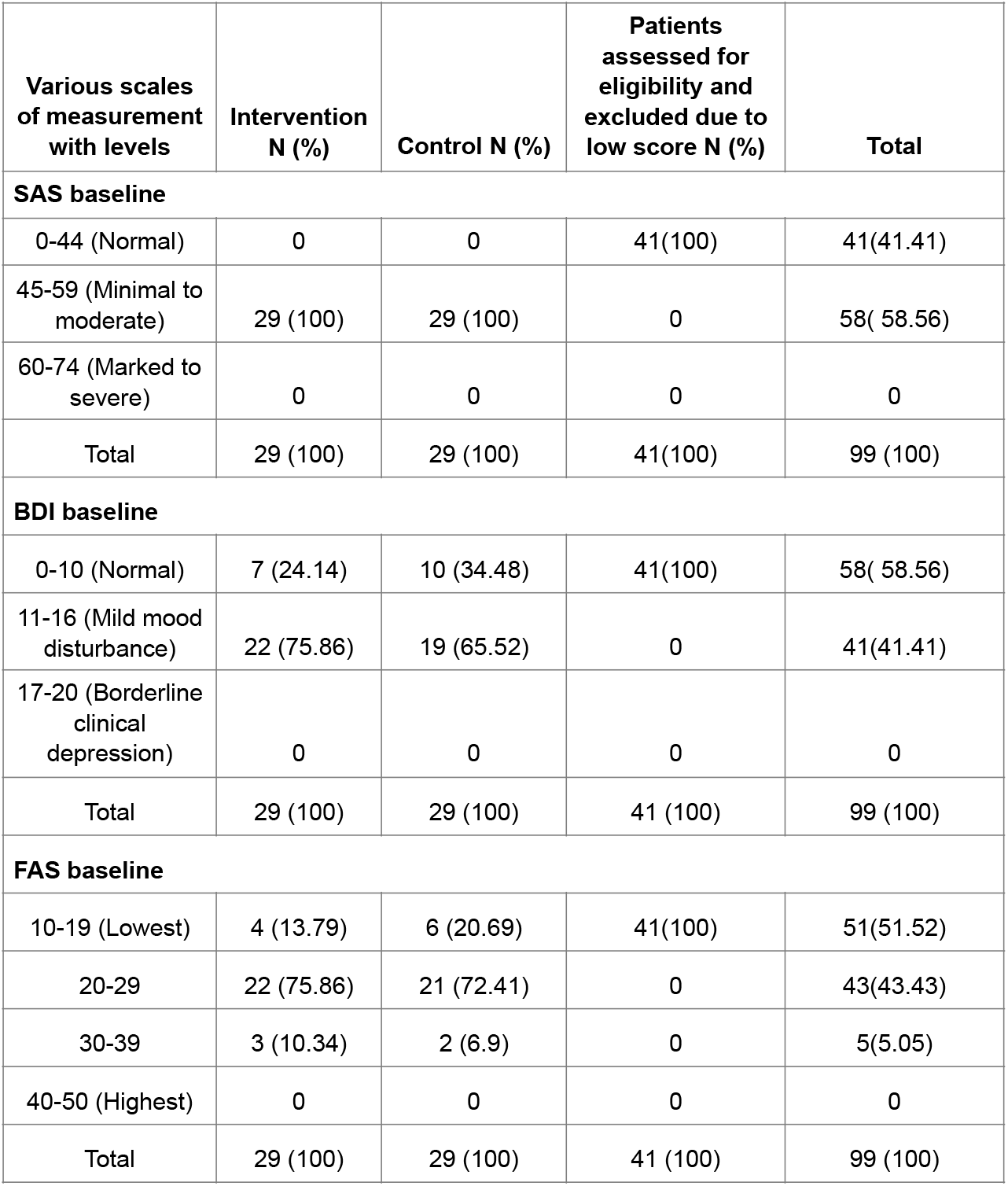
Measurement of the baseline level of anxiety, depression, and fatigue in breast cancer patients (N=58) *SAS-Zung Self-Rating Anxiety Scale, BDI-Beck Depression Inventory, FAS-Fatigue Assessment Scale*

When we include the patients, who were assessed for eligibility and excluded due to low scores, out of 99 patients, 58(58.56%) had Minimal to moderate levels of anxiety, 41(41.41%) had mild mood disturbance and 43(43.43%) had moderate level fatigue.

### 3.4 Comparison of mean scores of anxiety, depression, and fatigue in experimental and control groups (n = 58)

After two weeks of intervention in the experimental group, we measured the anxiety, depression, and fatigue scores in the experimental and control groups. No harm or unintended effects were noted in the intervention group. Table 4 and Figures 2-4 illustrate the comparison of mean anxiety, depression, and fatigue scores between the experimental and control groups.

**Table 4.** Comparison of Mean Anxiety, Depression, and Fatigue Scores between the Experimental and Control Groups. SD=Standard Deviation, P-value=Probability value, t= t-statistic

| Variable | Baseline score | After 2 weeks score | Paired t-test | Degree of freedom(df) | P-value |
| --- | --- | --- | --- | --- | --- |
| **Anxiety score** |  |  |  |  |  |
| **Intervention (Mean ± SD)** | 51.71 ± 2.89 | 38.52 ± 6.32 | t = 10.85 | 42.74 | 0.001 |
| **Control (Mean ± SD)** | 49.21 ± 2.44 | 49.14 ± 4.07 | t = 0.08 | 45.84 | 0.94 |
| **Depression score** |  |  |  |  |  |
| **Intervention (Mean ± SD)** | 11.48 ± 2.93 | 6.16 ± 2.98 | t = 7.09 | 59.98 | 0.001 |
| **Control (Mean ± SD)** | 10.66 ± 2.45 | 10.38 ± 2.46 | t = 0.43 | 56 | 0.67 |
| **Fatigue score** |  |  |  |  |  |
| **Intervention (Mean ± SD)** | 23.29 ± 4.12 | 16.88 ± 4.73 | t = 6.45 | 58.71 | ≤ 0.001 |
| **Control (Mean ± SD)** | 22.21 ± 4.82 | 22.34 ± 5.21 | t = -0.11 | 53.76 | 0.91 |

**Figure 2.**
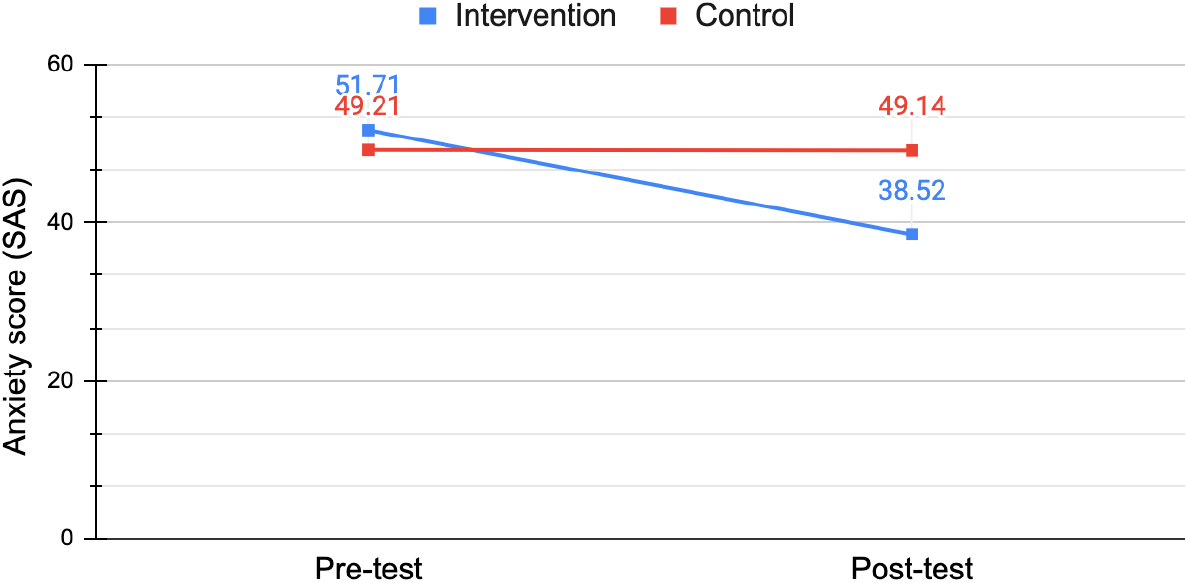
Mean SAS Scores Before and After the Intervention in Each Group

Anxiety in the intervention group decreased from 51.71 ± 2.89 to 38.52 ± 6.32 after 2 weeks of intervention (p ≤ 0.001). Anxiety in the control group also reduced from 49.21 ± 2.44 to 49.14 ± 4.07, but the difference is not statistically significant (p-value = 0.91). Postintervention, the anxiety scores in the intervention group (38.52 ± 6.32) were significantly lower (p-value ≤ 0.001) than those of the control group (49.14 ± 4.07) (Table 4, Figure 2).

Depression in the intervention group decreased from 11.48 ± 2.93 to 6.16 ± 2.98 after the intervention (p ≤ 0.001). Post-intervention, the depression scores in the intervention group (6.16 ± 2.98) were significantly low (p-value, ≤ 0.001) compared to the control group (10.38 ± 2.46) (Table 4, Figure 3).

**Figure 3.**
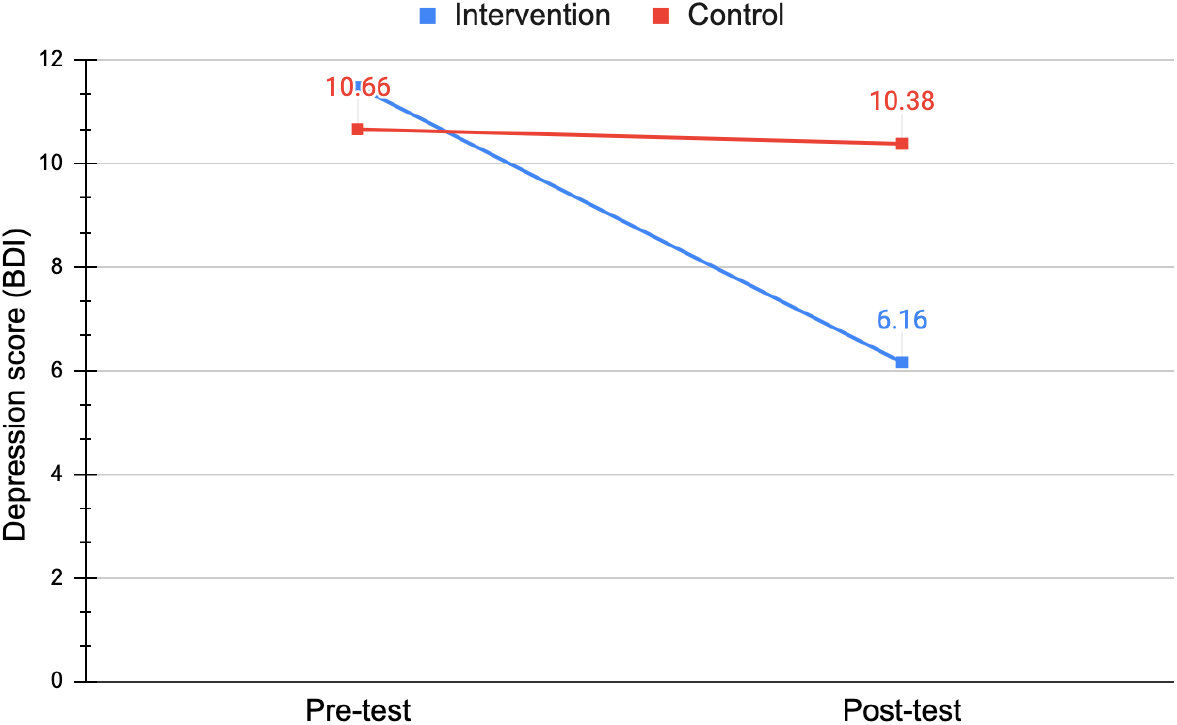
Mean Depression Scores Before and After the Intervention in Each Group.

Fatigue in the intervention group decreased from 23.29 ± 4.12 to 16.88 ± 4.73 after the intervention (p ≤ 0.001). Post-intervention, the fatigue scores in the intervention group (16.88 ± 4.73) were significantly lower (p-value ≤ 0.001) than those of the control group (22.34 ± 5.21) (Table 4, Figure 4).

**Figure 4.**
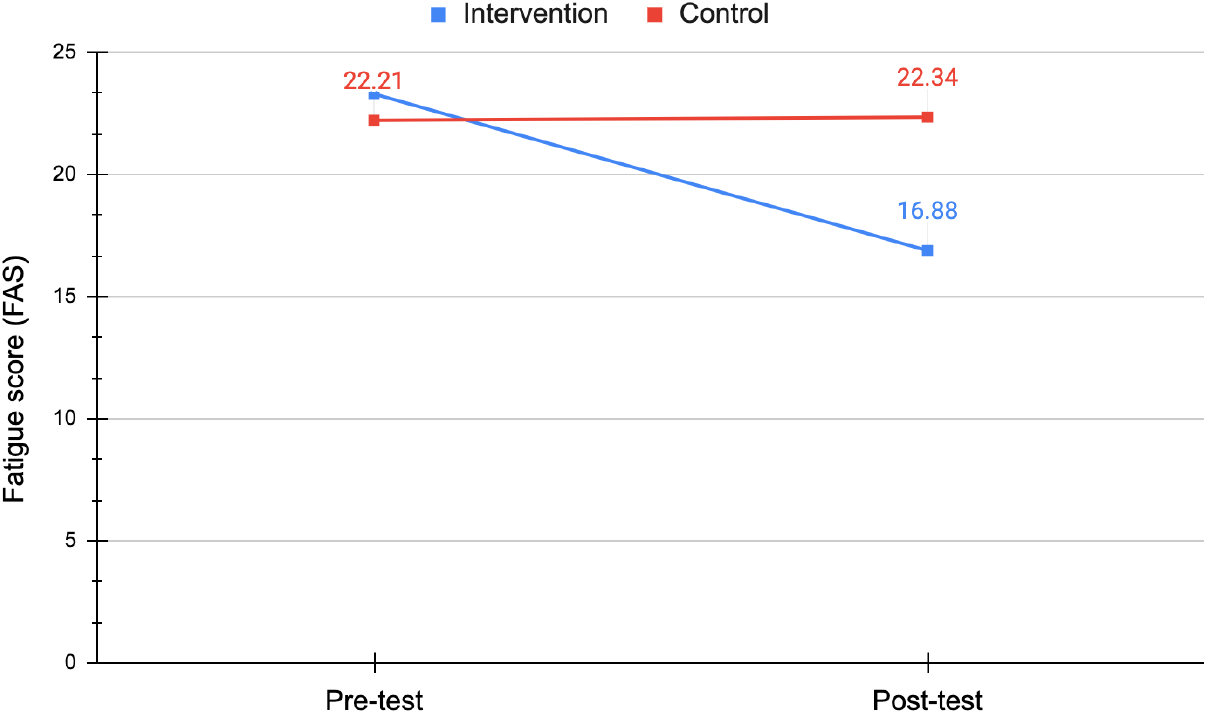
Mean Fatigue Scores Before and After the Intervention in Each Group

The average number of PMR exercises performed per day by the intervention group was 3.03. The average time taken to feel relaxed after PMR in minutes was 8.5 minutes, and the rate of relaxation they achieved in percentage was 80.4%, according to their feedback (Table 5).

**Table 5.**
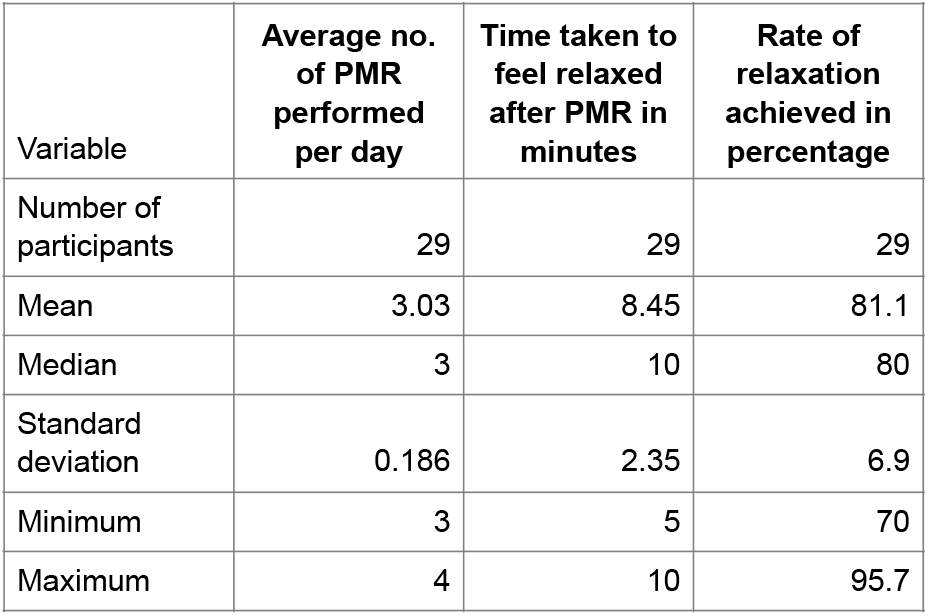
The data on home practice and patient feedback of the intervention group. *PMR=Progressive Muscle Relaxation*

| Variable | Average no. of PMR performed per day | Time taken to feel relaxed after PMR in minutes | Rate of relaxation achieved in percentage |
| --- | --- | --- | --- |
| Number of participants | 29 | 29 | 29 |
| Mean | 3.03 | 8.45 | 81.1 |
| Median | 3 | 10 | 80 |
| Standard deviation | 0.186 | 2.35 | 6.9 |
| Minimum | 3 | 5 | 70 |
| Maximum | 4 | 10 | 95.7 |

## 4 Discussion

In a study by Reyes-Gibby et al. (2015), fatigue was reported as one of the most prevalent symptoms among breast cancer patients undergoing chemotherapy, affecting nearly 40-50% of participants, which correlates with our finding of 43.43% of patients reporting moderate fatigue [22]. Furthermore, anxiety and mood disturbances are also commonly reported in breast cancer patients, with studies highlighting that up to 60% experience some form of anxiety during or after treatment [23] (Hamer et al., 2019). Our finding of 58.56% of patients experiencing minimal to moderate anxiety is in line with this data.

Furthermore, the work of Stark and House (2017) indicated that there is an underreporting of psychological distress (anxiety and mood disorders in particular) among breast cancer patients. Their studies found that about 40% of the patients developed mood dysfunction during treatment, which agrees with our finding of 41.41% of patients with mild mood disturbance [24].

The consistency of these findings across different studies suggests that it is crucial to initiate treatment of the psychological and physical symptoms toward the early stages to improve the psychosocial well-being of breast cancer patients.

The experimental group showed a significant reduction in anxiety (p ≤ 0.001), depression (p ≤ 0.001), and fatigue (p ≤ 0.001) from pre- to post-intervention, as determined by a paired t-test. This indicates a statistically significant decrease in these symptoms following the intervention. In contrast, no significant reductions were observed in the control group.

When comparing post-intervention scores between the two groups using a two-sample t-test, there were substantial differences in the mean scores of anxiety (p ≤ 0.001), depression (p ≤ 0.001), and fatigue (p ≤ 0.001). These findings confirm that the experimental group experienced a significantly greater reduction in anxiety, depression, and fatigue compared to the control group.

Similar results were reported in a study by Kaur et al., where participants in the experimental group showed a significant decrease in mean anxiety, depression, and fatigue scores (p ≤ 0.001 for all three variables) after 4½ weeks of radiotherapy or chemotherapy combined with Progressive Muscle Relaxation (PMR) and Guided Imagery (GI). Conversely, the control group experienced an increase in mean anxiety scores from 37.53 ± 5.10 to 39.77 ± 7.55 (p = 0.73), along with a significant rise in depression and fatigue scores (p ≤ 0.001) [6].

A randomized controlled trial by Charalambous, involving 256 patients undergoing chemotherapy for breast and prostate cancer, examined the effects of Progressive Muscle Relaxation (PMR) and Guided Imagery (GI) on reducing anxiety and depression. The study found that the intervention group experienced a decrease in mean anxiety scores after three weeks of intervention, while the control group showed a significant increase in anxiety scores from 39.47 ± 9.9 to 44.38 ± 7.6 [9].

Similarly, a study by Peterson, a randomized controlled trial focused on preventing anxiety and depression in gynecological cancer patients, produced comparable results. In this study, 53 patients were randomly assigned to either a control or intervention group. The intervention, which included counseling and relaxation sessions, led to a significant reduction in total hospital anxiety and depression scores, with notable decreases in the anxiety and moderate depression subscales (p = 0.001 and p = 0.02, respectively) [12].

León-Pizarro et al., in a randomized trial investigated the effect of training in relaxation and guided imagery techniques in improving psychological and quality-of-life indices for 66 gynecologic and breast brachytherapy patients. The intervention group experienced a statistically significant anxiety reduction (p = 0.008), depression (p = 0.03), and body discomfort (p = 0.04) compared to the control group [9, 25].

In a randomized controlled trial by Shu-Fen Chen et al., the experimental group demonstrated significant reductions in insomnia (-0.34 ± 0.83, p < 0.05), pain (-0.28 ± 0.58, p < 0.05), anxiety (-3.56 ± 2.94, p < 0.00), and depression (-2.38 ± 2.70, p < 0.00) between the pretest and posttest. When comparing the two groups, statistically significant differences were observed in overall symptom distress (B = 0.11, p < 0.05), insomnia (B = 0.50, p < 0.05), depression (B = 0.38, p < 0.05), and numbness in physical symptoms (B = 0.38, p < 0.05). Additionally, significant differences were found in anxiety (B = 3.08, p < 0.00) and depression (B = 1.86, p < 0.00) related to psychological distress [26].

### 4.1 Clinical Implications

Jacobson Progressive Muscle Relaxation (PMR) technique is a well-established method used in various medical and psychological contexts to alleviate stress and anxiety and promote relaxation. Based on our research on its application in breast cancer patients, we can draw upon its general clinical implications and potential benefits in this population:

PMR serves as a non-pharmacological intervention to address these psychological issues by inducing relaxation responses, reducing physiological arousal, and fostering a sense of calmness and well-being. By promoting relaxation and easing muscle tension, PMR reduces stress and improves sleep and pain reduction. Also, it enhances immunity by decreasing cortisol levels [9]. We could not prove it in this current study, further research is needed. Overall psychological well-being is the final assured outcome proved in our study. Therefore, we recommend that healthcare professionals integrate or suggest these techniques as adjuvant therapy for patients undergoing cancer treatment to help reduce anxiety and depression. Cancer patients often experience significant emotional distress even after treatment, highlighting the need for continued intervention beyond hospital discharge. However, this study did not assess the long-term effects of PMR. To ensure sustained implementation, it is crucial to enhance community networks for cognitive behavioral care and provide healthcare professionals with education in cognitive behavioral skills [9]. The intervention package can also be applied to other types of cancers and various disease conditions.

### 4.2 Limitations

The limitations of this study are that only hospitalized patients were used as participants, the sample size was small, and the study duration was short. Therefore, these findings may not be applicable to the general population. Also, the use of an open-label design in my study could lead to an exaggeration of the estimation of results compared with double-blind trials. It was difficult to mask the patients based on the intervention provided. So, there was difficulty in controlling the placebo effect. Even though patients reported that they adhered to the daily PMR exercise, I was not able to know whether they performed the exercise flawlessly each time and whether they could choose an external stimuli-free environment during the unsupervised sessions. Despite the limitations of the trial, the meticulous design and implementation of the study allow for the generalizability of the findings in this group of patients.

### 4.3 Future Research

Future research should include larger sample sizes and additional measures, such as biological markers (e.g., salivary cortisol), to improve measurement accuracy and enhance the generalizability of the findings. However, future studies can adopt a double-blind design and measure the long-term effects of these methods to provide more substantial evidence. Other nonpharmacological methods can also be compared with the JPMR technique for their effectiveness in relieving anxiety, depression, and fatigue.

## 5 Conclusions

In this study, the experimental group showed a statistically significant reduction in anxiety, depression, and fatigue levels (p ≤ 0.001 for all three variables) after two weeks of post-treatment, while no significant decrease was observed in the control group.

Patients undergoing cancer treatment often experience anxiety and depression. Incorporating daily relaxation techniques, such as Progressive Muscle Relaxation (PMR) and Guided Imagery (GI), can help alleviate overall symptom distress, anxiety, depression, and fatigue. These techniques are particularly beneficial for breast cancer patients during treatment. In this population, the combination of PMR and GI is effective in reducing anxiety, depression, and fatigue, and also preventing further progression.

Nonpharmacological therapy modalities are now recognized as valuable adjuncts to pharmacological therapy. In our study, the nonpharmacologic method was effective in the management of minimal to moderate anxiety, mild mood disturbances, and moderate-level fatigue in breast cancer patients. It also prevents the progression of anxiety, depression, and fatigue.

## Supporting information

Annexures

IEC file

CONSORT checklist

## Data Availability

All data produced in the present study are available upon reasonable request to the corresponding author.

## ACKNOWLEDGEMENT

I thank Dr. M. S. Kulkarni (Statistician, Dept. of Community Medicine, Goa Medical College) for his valuable statistical guidance for my research study. I thank Dr. Nitin Dhupdale (Lecturer, Dept. of Community Medicine, Goa Medical College) for encouraging my studies. I also thank all my faculty, colleagues, and friends, especially Dr. Winnie Antony (Jr. Consultant Psychiatry, Mental Health Center, Trivandrum), Dr. Ashish Shrivastava (Dept. of Psychiatry, Goa Medical College), S/N. Netra (Ward 106, Surgery, Goa Medical College), S/N. Shweta Desai (Ward 109, Surgery, Goa Medical College), Dr. Agasin Medemmel (JR, Surgery, Goa Medical College), and Dr. Gaurangi (JR, Pathology, Goa Medical College) for their valuable suggestions and positive criticisms of my research.

