## Supplementary material for "Effectiveness of a Relaxation Technique on Reducing Anxiety, Depression, and Fatigue among Women with Breast Cancer in Goa: A Randomized Controlled Trial": Annexures

### **Annexure A: Participant Information Sheet**

You are requested to participate in the below study, which will take place at your Surgery ward, Goa Medical College.

**Title of study**

Effectiveness of a Relaxation Technique on reducing Anxiety and Depression among Women with Breast Cancer in Goa: A Randomized Controlled Trial

**Purpose of the study**

Patients with cancer tend to have a lot of stress, especially when trying to cope with treatment and pain. We will measure your anxiety using the Zung Self-Rating Anxiety Scale (SAS), depression by the Beck Depression Interview II (BDI-II), and fatigue by fatigue assessment scale. You will be recruited to our study based on your baseline score if you consent. Then you will be randomly assigned to two groups; the undersigned doctor will teach one group relaxation techniques. The other group will be taught the same after two weeks when you come back for review.

The intervention entailed a combination of Progressive Muscle Relaxation (PMR), deep breathing, and Guided Imagery (GI) sessions. PMR is a technique of alternately tensing and relaxing muscle groups in sequence throughout the body to induce relaxation of mind and body. GI is defined as a cognitive process that utilizes the imagination to bring about positive mind/body responses that stimulate the senses. For the consistency of the exercises, an information booklet with pictures will be given. Daily reminders (text messages) will be given.

You will be again assessed for anxiety, depression, and fatigue on the same scale used before. Your sociodemographic data and case details will be collected. Vital signs will be recorded. Your feedback about the relaxation techniques will be collected at the end of the study. Psychological interventions can help you gain a better sense of control over distressing symptoms.

**Why are you included in this study?**

You are an adult woman ≥ 18 years of age with a new diagnosis of breast cancer who has undergone surgery as your primary mode of treatment.

**Procedure**

We will measure your anxiety using the Zung Self-Rating Anxiety Scale (SAS) ^13^, depression by the Beck Depression Interview II (BDI-II), and fatigue by fatigue assessment scale. Based on your baseline score you will be recruited to our study based on your baseline score if you consent. Then you will be randomly assigned to two groups; the undersigned doctor will teach one group relaxation techniques. The other group will be taught the same after two weeks when you come back for review.

The intervention entailed a combination of PMR, deep breathing, and GI sessions. PMR is a technique of alternately tensing and relaxing muscle groups in sequence throughout the body to induce relaxation of mind and body. GI is defined as a cognitive process that utilizes the imagination to bring about positive mind/body responses that stimulate the senses. For the consistency of the exercises, an information booklet with pictures will be given. Daily reminders (text messages) will be given.

You will be again assessed for anxiety, depression, and fatigue on the same scale used before. Your sociodemographic data and case details will be collected. Vital signs will be recorded. Your feedback about the relaxation techniques will be collected at the end of the study.

**Risks to you**

None

**Benefits to you**

Participating in this study will not provide you with a financial benefit.

You will be taught relaxation techniques either at the start or end of the study based on the randomization of the intervention and control groups.

Based on the results of this study, this interventional package will be recommended for implementation to reduce anxiety and depression in cancer patients at Goa Medical College.

**Confidentiality**

All data collected from you will be kept strictly confidential, and apart from the study investigators, no information will be given to anyone, including institutional authorities. Your name and identifying information will not be disclosed in any publication.

**Your right to withdraw**

The participation in this study is not compulsory. Even after you consent to the study, you are free to withdraw from it.

**Problems or questions**

You will be given a copy of this form. If you have any problems or questions regarding the study, please feel free to contact any of the investigators whose details are given below. You will also be asked to provide your contact details so the study organizers can contact you.

**Investigator’s details**

****************************

**In case of any ethical queries, please contact**

****************************

### **Annexure B: Consent Form For Participants**

I confirm that I have read and understood the information sheet for the above study and had the opportunity to ask questions.

I, _________________________ voluntarily agree to participate in this study. I understand that the study will involve:

● Collection of my personal and clinical details.

● Collection of my investigation reports.

● Assessment of my anxiety, depression, and fatigue levels using the Zung Self-Rating Anxiety Scale (SAS), Beck Depression Interview II (BDI-II), and Fatigue Assessment Scale (FAS).

● Learning of the intervention entailed a combination of relaxation techniques like PMR, deep breathing, and Guided Imagery sessions.

I understand that the information I provide will be used only to fulfill the above-stated objective. It will not be used in any harmful or discriminatory way against me.

I am also aware that I can withdraw from the study without stating any reasons for it, without my rights being affected.

I agree that the data or results that arise from the study can be used for a scientific purpose, and there are no monetary benefits.

Signature of the participant

Name of the participant

Signature of the investigator

**Investigator’s details**

****************************

Date:

Time:

### **Annexure C: Case Record Form (CRF)**

| Serial  Number | Question | Possible answers |
| --- | --- | --- |
| 1 | Date |  |
| 2 | Hospital Number |  |
| 3 | Ward No. |  |
| 4 | Name |  |
| 5 | Age |  |
| 6 | Sex | ❏ Male  ❏ Female |
| 7 | Address and phone number |  |
| 8 | Religion |  |
| 9 | Habitat | Urban ❏ Rural |
| 10 | Marital status | ❏ Married  ❏ Widowed/Divorced  ❏ Single |
| 11 | Education | ❏ Illiterate  ❏ Primary  ❏ Secondary  ❏ Higher Secondary  ❏ Graduate |
| 12 | Occupation | ❏ Unemployed  ❏ Employed |
| 13 | Income | ❏ Upper class (7533 and above)  ❏ Upper middle (3766-7532)  ❏ Lower middle (2260-3765)  ❏ Upper lower (1130-2259)  ❏ Lower (1129 and Below) |
| 14 | Patient perception of support level  (score 0 to 5) |  |
| 15 | Anesthetic coding (ASA score) |  |
| 16 | Surgical procedures undertaken |  |
| 17 | Length of surgery (min) |  |
| 18 | Post-operative complications |  |
| 19 | Postoperative hospital stay (days) |  |
| 20 | Adjuvant therapy required |  |
| 21 | Tumor type/Stage/Grade |  |
| 22 | Presenting symptom  presentation (days) |  |
| 23 | Time from onset  of symptoms to medical |  |
| 24 | Time from onset of  symptoms to diagnosis (days) |  |
| 25 | Time from medical  presentation to treatment (days) |  |
| 26 | Family history of breast cancer |  |
| 27 | Obstetric history |  |
| 28 | USG (BIRADS) |  |
| 29 | Vitals  BP, HR |  |

### **Annexure D: Zung Self-Rating Anxiety Scale (SAS)**

For each item below, please place a checkmark (✔️) in the column which best describes how often you felt or behaved this way during the past several days. Bring the completed form with you to the office for scoring and assessment during your office visit. (Source: William W.K. Zung. A rating instrument for anxiety disorders. Psychosomatics. 1971)

| S.No. | Place a checkmark (✔️) in the correct column. | A little of  the time | Some of  the time | A good part of  the time | Most of  the time |
| --- | --- | --- | --- | --- | --- |
| 1 | I feel more nervous and anxious than usual. |  |  |  |  |
| 2 | I feel afraid for no reason at all. |  |  |  |  |
| 3 | I get upset easily or feel panicky. |  |  |  |  |
| 4 | I feel like I'm falling apart and going to pieces. |  |  |  |  |
| 5 | I feel that everything is alright and nothing bad will happen. |  |  |  |  |
| 6 | My arms and legs shake and tremble. |  |  |  |  |
| 7 | I am bothered by headaches, neck, and back pain. |  |  |  |  |
| 8 | I feel weak and get tired easily. |  |  |  |  |
| 9 | I feel calm and can sit still easily. |  |  |  |  |
| 10 | I can feel my heart beating fast. |  |  |  |  |
| 11 | I am bothered by dizzy spells. |  |  |  |  |
| 12 | I have fainting spells or feel like it. |  |  |  |  |
| 13 | I can breathe in and out easily. |  |  |  |  |
| 14 | I get feelings of numbness and tingling in my fingers & toes. |  |  |  |  |
| 15 | I am bothered by stomach aches or  indigestion. |  |  |  |  |
| 16 | I have to empty my bladder often. |  |  |  |  |
| 17 | My hands are usually dry and warm. |  |  |  |  |
| 18 | My face gets hot and blushes. |  |  |  |  |
| 19 | I fall asleep easily and get a good night's rest. |  |  |  |  |
| 20 | I have nightmares. |  |  |  |  |

### **Annexure E: Beck's Depression Inventory**

This depression inventory can be self-scored. The scoring scale is at the end of the questionnaire.

| S.No. | Score |  |
| --- | --- | --- |
| 1 | 0 | I do not feel sad. |
|  | 1 | I feel sad |
|  | 2 | I am sad all the time, and I can't snap out of it. |
|  | 3 | I am so sad and unhappy that I can't stand it. |
| 2 | 0 | I am not particularly discouraged about the future. |
|  | 1 | I feel discouraged about the future. |
|  | 2 | I feel I have nothing to look forward to. |
|  | 3 | I feel the future is hopeless and that things cannot improve. |
| 3 | 0 | I do not feel like a failure. |
|  | 1 | I feel I have failed more than the average person. |
|  | 2 | As I look back on my life, all I can see is a lot of failures. |
|  | 3 | I feel I am a complete failure as a person. |
| 4 | 0 | I get as much satisfaction out of things as I used to. |
|  | 1 | I don't enjoy things the way I used to. |
|  | 2 | I don't get real satisfaction out of anything anymore. |
|  | 3 | I am dissatisfied or bored with everything. |
| 5 | 0 | I don't feel particularly guilty |
|  | 1 | I feel guilty a good part of the time. |
|  | 2 | I feel quite guilty most of the time. |
|  | 3 | I feel guilty all of the time. |
| 6 | 0 | I don't feel I am being punished. |
|  | 1 | I feel I may be punished. |
|  | 2 | I expect to be punished. |
|  | 3 | I feel I am being punished. |
| 7 | 0 | I don't feel disappointed in myself. |
|  | 1 | I am disappointed in myself. |
|  | 2 | I am disgusted with myself. |
|  | 3 | I hate myself. |
| 8 | 0 | I don't feel I am any worse than anybody else. |
|  | 1 | I am critical of myself for my weaknesses or mistakes. |
|  | 2 | I blame myself all the time for my faults. |
|  | 3 | I blame myself for everything bad that happens. |
| 9 | 0 | I don't have any thoughts of killing myself. |
|  | 1 | I have thoughts of killing myself, but I would not carry them out. |
|  | 2 | I would like to kill myself. |
|  | 3 | I would kill myself if I had the chance. |
| 10 | 0 | I don't cry any more than usual. |
|  | 1 | I cry more now than I used to. |
|  | 2 | I cry all the time now. |
|  | 3 | I used to be able to cry, but now I can't cry even though I want to. |
| 11 | 0 | I am no more irritated by things than I ever was. |
|  | 1 | I am slightly more irritated now than usual. |
|  | 2 | I am quite annoyed or irritated a good deal of the time. |
|  | 3 | I feel irritated all the time. |
| 12 | 0 | I have not lost interest in other people. |
|  | 1 | I am less interested in other people than I used to be. |
|  | 2 | I have lost most of my interest in other people. |
|  | 3 | I have lost all of my interest in other people. |
| 13 | 0 | I make decisions about as well as I ever could. |
|  | 1 | I put off making decisions more than I used to. |
|  | 2 | I have greater difficulty in making decisions than I used to. |
|  | 3 | I can't make decisions at all anymore. |
| 14 | 0 | I don't feel that I look any worse than I used to. |
|  | 1 | I am worried that I am looking old or unattractive. |
|  | 2 | I feel there are permanent changes in my appearance that make me look unattractive |
|  | 3 | I believe that I look ugly. |
| 15 | 0 | I can work about as well as before. |
|  | 1 | It takes an extra effort to get started at doing something. |
|  | 2 | I have to push myself very hard to do anything. |
|  | 3 | I can't do any work at all. |
| 16 | 0 | I can sleep as well as usual. |
|  | 1 | I don't sleep as well as I used to. |
|  | 2 | I wake up 1-2 hours earlier than usual and find it hard to get back to sleep |
|  | 3 | I wake up several hours earlier than I used to and cannot get back to sleep. |
| 17 | 0 | I don't get more tired than usual. |
|  | 1 | I get tired more easily than I used to. |
|  | 2 | I get tired from doing almost anything. |
|  | 3 | I am too tired to do anything. |
| 18 | 0 | My appetite is no worse than usual. |
|  | 1 | My appetite is not as good as it used to be. |
|  | 2 | My appetite is much worse now. |
|  | 3 | I have no appetite at all anymore. |
| 19 | 0 | I haven't lost much weight, if any, lately. |
|  | 1 | I have lost more than five pounds. |
|  | 2 | I have lost more than ten pounds. |
|  | 3 | I have lost more than fifteen pounds. |
| 20 | 0 | I am no more worried about my health than usual. |
|  | 1 | I am worried about physical problems like aches, pains, upset stomach, Or constipation. |
|  | 2 | I am very worried about physical problems and it's hard to think of much else. |
|  | 3 | I am so worried about the physical problems that I cannot think of anything else. |
| 21 | 0 | I have not noticed any recent change in my interest in sex. |
|  | 1 | I am less interested in sex than I used to be. |
|  | 2 | I have almost no interest in sex. |
|  | 3 | I have lost interest in sex completely. |

INTERPRETING THE BECK DEPRESSION INVENTORY

Now that you have completed the questionnaire, add up the score for each of the twenty-one questions by counting the number to the right of each question you marked. The highest possible total for the whole test would be sixty-three. This would mean you circled number three on all twenty-one questions. Since the lowest possible score for each question is zero, the lowest possible score for the test would be zero. This would mean you circle zero on each question. You can evaluate your depression according to the Table below.

Total Score____________________Levels of Depression

1-10____________________These ups and downs are normal

11-16___________________ Mild mood disturbance

17-20___________________Borderline clinical depression

21-30___________________Moderate depression

31-40___________________Severe depression

over 40__________________Extreme depression

### **Annexure F: Fatigue Assessment Scale (FAS)**

The following 10 statements refer to how you usually feel. For each statement you can choose one out of five answer categories, varying from never to always.1 = never, 2 = sometimes; 3 = regularly; 4 = often; and 5 = always.

| S/N |  | Never | Sometimes | Regularly | Often | Always |
| --- | --- | --- | --- | --- | --- | --- |
| 1 | I am bothered by fatigue (WHOQOL) | 1 | 2 | 3 | 4 | 5 |
| 2 | I get tired very quickly (CIS) | 1 | 2 | 3 | 4 | 5 |
| 3 | I don't do much during the day (CIS) | 1 | 2 | 3 | 4 | 5 |
| 4 | I have enough energy for everyday life (WHOQOL) | 1 | 2 | 3 | 4 | 5 |
| 5 | Physically, I feel exhausted (CIS) | 1 | 2 | 3 | 4 | 5 |
| 6 | I have problems starting things (FS) | 1 | 2 | 3 | 4 | 5 |
| 7 | I have problems thinking clearly (FS) | 1 | 2 | 3 | 4 | 5 |
| 8 | I feel no desire to do anything (CIS) | 1 | 2 | 3 | 4 | 5 |
| 9 | Mentally, I feel exhausted | 1 | 2 | 3 | 4 | 5 |
| 10 | When I am doing something, I can concentrate quite well (CIS) | 1 | 2 | 3 | 4 | 5 |

Note: The abbreviations after the items indicate the scale from which the items have been abstracted. CIS - Checklist Individual Strength; WHOQOL - World Health Organization Quality of Life assessment instrument; FS - Fatigue Scale. Items 4 and 10 require reversed scoring. The scale score is calculated

by summing all items. Total scores can range from 10, indicating the lowest level of fatigue, to 50, denoting the highest.

### **Annexure G: Jacobson Progressive Muscle Relaxation (PMR) Technique**

General Instruction (Before and During Muscle Relaxation Exercise):

1. Sit or lie down in a comfortable position.

2. During the part of the exercise cycle, tense the muscle tightly and hold for a slow count of 5 seconds. (Repeat silently 1001, 1002, 1003,…)

3. During the relation part of the exercise cycle, relax the muscle quickly, and completely. Let your mind relax and appreciate how relaxed the muscle is feeling for 10 seconds.

4. Try to keep all other muscles relaxed as you exercise a specific muscle group.

5. As you exercise from head to toe................ Observe changes like tightness and the development of light and soothing sensations.

6. Deep Breathing: Relax by taking three deep breaths inhaling through the nose and exhaling through the mouth after each step. This consists of breathing through the belly and not the chest more slowly and deeply. You may say “in two-three four” and “out two three four”.

7. Guided Imagery: After taking a few deep breaths, let your mind wander into a special place that you associate with relaxation. For some people, this may be a beach. For others, somewhere out in nature. It may even be in the comfort of your own home. Take a moment to bring this place to mind. Then visualize yourself there seeing the colors and shape, taking in smells and feelings and sensations. Allow yourself to become involved in experiencing all these images. What are the colors? Smells? Is there a breeze or warm ray of sunlight in the trees or a soft murmur of a stream or a crash of waves on a beach? Bring yourself to this relaxing place, and feel physically and emotionally relaxed.

8. Now make your body completely loose.............light.............. and free

9. Let us begin your exercise.

| S/N | PROCEDURE OF JACOBSON'S PROGRESSIVE MUSCLE RELAXATION TECHNIQUE | | Tensing  Time | Relaxation Time |
| --- | --- | --- | --- | --- |
| 1 | Hands | |  |  |
|  | a) | Clench each fist separately (right & left), feel the tension in the fist and forearm respectively for 5 seconds  Release the fist, relax and feel relaxation for 10 seconds | 5 sec | 10 sec |
| 2 | Arms | |  |  |
|  | a) | Bend each arm separately (right & left) up at the elbow and tense the biceps, keeping the hand relaxed, feel the tension for 5 seconds.  Release the arm, relax and feel relaxation for 10 seconds | 5 sec | 10 sec |
|  | b) | Straighten the arm separately (right & left) and tense the triceps leaving the lower arms supported by the chair with the hands relaxed, feel tense for 5 seconds. Relax and feel relaxation for 10 seconds | 5 sec | 10 sec |
|  | Facial Muscles | |  |  |
|  | a) | Wrinkle your forehead; try to make your eyebrows touch your hairline which produces tension, feel the tension for 5 seconds. Release the eyebrows and relax for 10 seconds. | 5 sec | 10 sec |
|  | b) | Close your eyes and screw the muscles, around the eyes for 5 seconds. Release, relax, and feel relaxed for 10 seconds. | 5 sec | 10 sec |
|  | c) | Tense the jaw by biting the teeth together, feel the tension in the jaw muscles for 5 seconds. Release, relax and feel relaxed for 10 seconds. | 5 sec | 10 sec |
|  | d) | Press the tongue hard and flat against the roof of the mouth with lips closed, notice tension in the throat, and feel it for 5 seconds. Release, relax and feel relaxed for 10 seconds. | 5 sec | 10 sec |
| 3 | Neck & shoulder | |  |  |
|  | a) | Push the head back as far as it will go (against a chair), feel the tension for 5 seconds. Bring the head to its position, relax, and feel relaxed for 10 seconds. | 5 sec | 10 sec |
|  | b) Bring the head down and press the chin down onto the chest for 5 seconds. Bring the head to its  position, relax, and feel the relaxation for 10  seconds.  c) Tense shoulders by tightening and shrinking shoulders (Shrug your shoulders up to your ears) feel the tension for 5 seconds.  Release, relax and feel relaxed for 10 seconds. | | 5 sec | 10 sec |
| 4 | Chest |  |  |  |
|  | a) Take a deep breath, filling the lungs, hold the breath for a few seconds, and passively exhale. Relax and feel relaxed for 10 seconds. | | 5 sec | 10 sec |
|  |  | Relax and feel relaxed for 10 seconds. |  |  |
| 5 | Stomach. | |  |  |
|  | a) | Pull in the stomach and tense the stomach muscle for 5 seconds.  Release the stomach, relax and feel relaxation for 10 seconds | 5 sec | 10 sec |
| 6 | Back | |  |  |
|  | a) | Arch your back away from the chair and feel tension for 5 seconds.  Relax and feel relaxed for 10 seconds. | 5 sec | 10 sec |
| 7 | Thighs & Buttocks | |  |  |
| 8 | a) Tense both thigh muscles and buttocks by squeezing muscles together and feel tensing for 5 seconds.  Release the muscles, relax, and feel relaxed for 10 seconds. Lower Legs  a) Point toes towards your head, producing tension in calf muscles; feel tense for 5 seconds. Relax and feel relaxed for 10 seconds.  b) Point the toes away from the head, feel the tension for 5 seconds. Relax and feel relaxation for 10 seconds | | 5 sec  5 sec  5 sec | 10 sec  10 sec  10 sec |
| 9 | Toes | |  |  |
|  | a) | Curl your toes in tightly to feel the tensing muscles of the foot, of both feet. Hold this for a few moments before releasing the tension and relaxing the feet. Relax and feel relaxed for 10 seconds. | 5 sec | 10 sec |
| 10 | After Exercises | |  |  |
|  | a) | Relax the whole body completely. |  | 2min  relaxation |
|  | b) | Keep your eyes closed and let yourself remain in a relaxed position. |  |  |
|  | c) | Open your eyes and enjoy renewed energy, feel relaxed and refreshed. |  |  |
|  | d) | Sit up, stretch, and stand up slowly. |  |  |

You can do this in any place or situation. It can be done partially, also involving a few muscles or a single muscle group. You can end the exercise with a few deep breaths and guided imagery to feel complete relaxation.

| Days | No of Relaxation  technique performed per day | How much time does it take you to get relaxed, during exercise? ( in  minutes) | Rate your relaxation on a scale of 1 to 100 |
| --- | --- | --- | --- |
| 1 |  |  |  |
| 2 |  |  |  |
| 3 |  |  |  |
| 4 |  |  |  |
| 5 |  |  |  |
| 6 |  |  |  |
| 7 |  |  |  |
| 8 |  |  |  |
| 9 |  |  |  |
| 10 |  |  |  |
| 11 |  |  |  |
| 12 |  |  |  |
| 13 |  |  |  |
| 14 |  |  |  |
| 15. |  |  |  |
| Please write your valuable Feedback about the relaxation technique: | | | |
