## Supplementary material for "Effectiveness of a Relaxation Technique on Reducing Anxiety, Depression, and Fatigue among Women with Breast Cancer in Goa: A Randomized Controlled Trial": IEC file

### GOA MEDICAL COLLEGE

#### INSTITUITIONAL ETHICS COMMITTEE

Office: Dept of Pharmacology, Goa Medical College, Bambolim Complex, Goa 403202

**Chairman**

Dr Philomina DSouza

Cell: 7769043243

**Member Secretary**

Dr Amey Kamat

Cell: 9822751356

**Members**

Dr Viraj Khandeparkar

Cell: 9421138116

Dr. Noel Menezes

Cell: 9922981002

Dr Rakhee Ghodge

Cell: 9822150305

Dr Jagadish Bhat

Cell: 9673217939

Dr. Pandarinath Audi

Mrs Sandhya Ram

Dr.Nandita De Souza

Mr Pradip J. Mhamai Kamat

Dr Anasuya Ganguly

Ms Vedita Hegde Desai

Dr Lois J Samuel

Dr Amita Kamat Kenkre

To,

Date: 26/12/20

Dr. Dhanya Jose

JR-PSM Department

GMC

Sub: - Approval of the study documents submitted by you.

Madam,

This is to inform you that IEC met on 24 Dec 2020 at 2 pm and reviewed an discussed your study documents on

**"Effectiveness of a Relaxation Technique on Reducing Anxiety and Depression among Women with Selected Cancers in Goa: A Randomized Controlled Trial."**

The study has been approved by the committee in its present form for its entire duration.

Any changes in the protocol have to be informed to the undersigned at the earliest.

Yours Sincerely,

Dr Amey Kamat

Member Secretary IEC

### GOA MEDICAL COLLEGE

#### INSTITUTIONAL ETHICS COMMITTEE

Office: Dept. of Pharmacology, Goa Medical College, Bambolim Complex, Goa 403202 . **Registration No. ECR/83/Inst/GOA/2013/RR-20**

**Chairperson**
Dr.Anasuya Ganguly
Cell: 9370192340
**Member Secretary**
Dr (Mrs) Lois J. Samuel
Cell: 9420165477
**Members**
Dr. Jagadish Cacodkar
Cell: 9823928743
Dr. Carlos Noel Menezes
Cell: 9822981002
Dr. Pandarinath Audi
Cell: 9561251420
(Mrs). Vedita Hegde Desai
Cell: 9823472942
Dr. Amita Kamat
Cell: 9823199347
Dr. Viraj Khandeparkar
Cell: 9421138116
Dr Sandhya Ram
Cell: 9823994865
Dr. Nandita De Souza
Cell: 9422634356
Mr Pradip Mhamai Kamat
Cell: 9422445655

To
Dr. Dhanya Jose
Junior Resident
Department of PSM
Goa Medical College
Bambolim- Goa.

17/05/2022

Sub:-Approval of the study submitted by you.

Madam,

This is to inform you that IEC met on 13/05/2022 at 3.00 pm and reviewed your study documents.

**Title – Effectiveness of a relaxation technique on reducing anxiety and depression among women with breast cancer in Goa : A Randomized Controlled Trial.**

The study has been approved in its present form.

Any changes in the protocol have to be informed to the undersigned at the earliest.

**Reference code: GMCIEC/2022/62** (for further correspondence)

Yours sincerely

Lois James Samuel

Dr (Mrs) Lois James Samuel

Member Secretary IEC

MEMBER SECRETARY
Institutional Ethics Committee
Goa Medical College

#### Clinical Trial Details (PDF Generation Date :- Fri, 11 Nov 2022 05:43:49 GMT)

| | |
|---|---|
| **CTRI Number** | CTRI/2021/02/030996 [Registered on: 04/02/2021] - **Trial Registered Prospectively** |
| **Last Modified On** | 11/11/2022 |
| **Post Graduate Thesis** | Yes |
| **Type of Trial** | Interventional |
| **Type of Study** | Other (Specify) [Psychological Intervention] |
| **Study Design** | Randomized, Parallel Group Trial |
| **Public Title of Study** | A study to find out effect of a Relaxation Exercise on Reducing Anxiety and Depression among Women with Breast and Reproductive organ Cancers in Goa |
| **Scientific Title of Study** | Effectiveness of a Relaxation Technique on Reducing Anxiety and Depression among Women with Breast Cancer in Goa: A Randomized Controlled Trial |

**Secondary IDs if Any**

| Secondary ID | Identifier |
|---|---|
| NIL | NIL |

**Details of Principal Investigator or overall Trial Coordinator (multi-center study)**

| Details of Principal Investigator | |
|---|---|
| **Name** | Dr Dhanya Jose |
| **Designation** | Junior Resident |
| **Affiliation** | Goa Medical College |
| **Address** | Department of Preventive and Social Medicine, Goa Medical College, Bambolim<br>North Goa<br>GOA<br>403202<br>India |
| **Phone** | 9747773075 |
| **Fax** | |
| **Email** | |

**Details Contact Person (Scientific Query)**

| Details Contact Person (Scientific Query) | |
|---|---|
| **Name** | Dr Jagadish Cacodcar |
| **Designation** | Professor and HOD |
| **Affiliation** | Goa Medical College |
| **Address** | Department of Preventive and Social Medicine, Goa Medical College, Bambolim<br>North Goa<br>GOA<br>403202<br>India |
| **Phone** | 9823928743 |
| **Fax** | |
| **Email** | |

**Details Contact Person (Public Query)**

| Details Contact Person (Public Query) | |
|---|---|
| **Name** | Dr Dhanya Jose |
| **Designation** | Junior Resident |
| **Affiliation** | Goa Medical College |
| **Address** | Department of Preventive and Social Medicine, Goa Medical College, Bambolim<br>North Goa<br>GOA<br>403202<br>India |

| | |
|---|---|
| **Phone** | 9747773075 |
| **Fax** | |
| **Email** | |

**Source of Monetary or Material Support**

| Source of Monetary or Material Support |
|---|
| > Goa Medical College, Bambolim-403202 |

**Primary Sponsor**

| Primary Sponsor Details | |
|---|---|
| **Name** | Dr Dhanya Jose |
| **Address** | Junior Resident Department of Preventive and Social Medicine Goa Medical College Bambolim-403202 |
| **Type of Sponsor** | Other [Self] |

**Details of Secondary Sponsor**

| Name | Address |
|---|---|
| NIL | NIL |

**Countries of Recruitment**

| List of Countries |
|---|
| India |

**Sites of Study**

| Name of Principal Investigator | Name of Site | Site Address | Phone/Fax/Email |
|---|---|---|---|
| Dr Dhanya Jose | Demonstration Room, Ward-106,109,129 | Female Surgical Ward-106,First Floor; Female Surgical Ward-109, Second Floor; gynecology Ward-129, Third Floor; Goa Medical College Hospital, Bambolim-403202<br>North Goa<br>GOA | 9747773075<br><br> |

**Details of Ethics Committee**

| Name of Committee | Approval Status | Date of Approval | Is Independent Ethics Committee? |
|---|---|---|---|
| Institutional Ethics Committee | Approved | 26/12/2020 | No |
| Institutional Ethics Committee | Approved | 17/05/2022 | No |

**Regulatory Clearance Status from DCGI**

| Status | Date |
|---|---|
| Not Applicable | No Date Specified |

**Health Condition / Problems Studied**

| Health Type | Condition |
|---|---|
| Patients | Malignant neoplasm of breast |
| Patients | Malignant neoplasms of female genital organs |

**Intervention / Comparator Agent**

| Type | Name | Details |
|---|---|---|
| Intervention | The intervention entails a 20 minutes relaxation session of Progressive Muscle Relaxation (PMR) with Deep Breathing and Guided Imagery (GI) sessions. | Progressive Muscle Relaxation (PMR) is a technique of alternately tensing and relaxing muscle groups in sequence throughout the body to induce relaxation of mind and body. Guided Imagery (GI) is defined as a cognitive process that utilizes the imagination to bring about positive mind/body responses that stimulate the senses. Deep breathing consists of breathing through |

| | | |
|---|---|---|
| | | the belly and not the chest more slowly and deeply. Deep breathing lowers your heart rate, your blood pressure, increases your energy level and decreases muscle tension and pain. |
| Comparator Agent | Standard cancer treatment and care followed at Goa Medical College | Currently no additional measures to measure and reduce anxiety and depression among cancer women. So control group will get only the Standard cancer treatment and care followed at Goa Medical College. |

**Inclusion Criteria**

| Inclusion Criteria | |
|---|---|
| **Age From** | 18.00 Year(s) |
| **Age To** | 65.00 Year(s) |
| **Gender** | Female |
| **Details** | 1) Adult women of age between 18 and 65 years with a new diagnosis of cancer, who have undergone surgery for Ca breast, Ca Cervix, Ca ovary, or Ca endometrium as their primary mode of treatment.<br/> 2)Those who are experiencing anxiety or depression (Zung anxiety scale score; SAA>44, Beck Depression Inventory; BDI>10, are not receiving medication for anxiety or depression.<br/> 3)Those who do not have cognitive impairment.<br/> |

**Exclusion Criteria**

| Exclusion Criteria | |
|---|---|
| **Details** | 1)Patients require medication for anxiety or depression during the study period.<br>2)Those who are not consenting to the study. |

| | |
|---|---|
| **Method of Generating Random Sequence** | Computer generated randomization |
| **Method of Concealment** | Sequentially numbered, sealed, opaque envelopes |
| **Blinding/Masking** | Open Label |

**Primary Outcome**

| Outcome | Timepoints |
|---|---|
| The primary outcome will be the difference between baseline and follow-up scores of anxiety, depression, and fatigue scales. | Both groups will be assessed at baseline and the end of the two-week intervention period for their anxiety, depression and fatigue levels. |

**Secondary Outcome**

| Outcome | Timepoints |
|---|---|
| Secondary outcomes include the scores of anxiety, depression, and fatigue scales and the patients overall impression of the intervention. | Both groups will be assessed at baseline and the end of the two-week intervention period for their anxiety, depression and fatigue levels. Patients feedback about the relaxation techniques will be collected at the end of the study. |

| | |
|---|---|
| **Target Sample Size** | **Total Sample Size=**60<br>**Sample Size from India=**60<br>**Final Enrollment numbers achieved (Total)=**0<br>**Final Enrollment numbers achieved (India)=**60 |
| **Phase of Trial** | N/A |
| **Date of First Enrollment (India)** | 08/02/2021 |
| **Date of First Enrollment (Global)** | No Date Specified |

| | |
|---|---|
| **Estimated Duration of Trial** | **Years=**1<br>**Months=**0<br>**Days=**0 |
| **Recruitment Status of Trial (Global)** | Not Applicable |
| **Recruitment Status of Trial (India)** | Completed |
| **Publication Details** | After completing the trial, the results will be published in peer reviewed indexed journal. |
| **Brief Summary** | **Objective:**<br>• Examine the effectiveness of a relaxation technique on reducing anxiety and depression in women with breast cancer<br>• Measure the level of anxiety, depression, and fatigue among women with breast cancer<br><br>**Design:** Randomised Controlled Trial<br><br>**Participants:** 60 female patients with breast cancer.<br><br>**Setting:** Surgery wards at Goa Medical College<br><br>**Methods:** The patient will be provided with information sheets in the postoperative period and will be invited to participate in the study. After getting the consent, we will measure the patient’s anxiety using the Zung Self-Rating Anxiety Scale (SAS), depression by the Beck Depression Interview II (BDI-II), and fatigue by fatigue assessment scale. Based on the baseline score, patients will be recruited for our study. Then randomly assigned to two groups, the intervention group will be taught relaxation techniques by the principal investigator. The control group will be taught the same after 2 weeks when they come back for review.<br><br>The intervention entails a 20 minutes relaxation session of Progressive Muscle Relaxation (PMR) with deep breathing and Guided Imagery (GI) sessions. PMR is a technique of alternately tensing and relaxing muscle groups in sequence throughout the body to induce relaxation of mind and body. GI is defined as a cognitive process that utilizes the imagination to bring about positive mind/body responses that stimulate the senses. For the consistency of the exercises, the information booklet with pictures will be given. Reminders (text messages) will be given.<br><br>The participant will be again assessed for anxiety, depression, and fatigue after 2 weeks on the same scale used before. The sociodemographic data and case details will be collected. Vital signs will be recorded. The participants’ feedback |

about the relaxation techniques will be collected at the end of the study.

**Outcome Assessment:** The primary outcome will be the difference between baseline and follow-up scores on anxiety, depression, and fatigue scales. Secondary outcomes include the scores of anxiety, depression, and fatigue scales and the patient's overall impression of the intervention.

**Data analysis:** The data will be entered using Google Sheets and analyzed using R program software. P-value < 0.05 will be taken as a level of significance. Student t-test and chi-square test will be used for statistical analysis. For a given scale, the individual difference between the baseline and the at the end of the intervention will be calculated using the paired t-test, and the mean difference between the two groups will be calculated by unpaired t-test.
